# Sex differences in incidence and mortality of bloodstream infections. Results from the population-based HUNT study in Norway

**DOI:** 10.1101/2021.08.24.21262516

**Authors:** Randi Marie Mohus, Lise T. Gustad, Anne Sofie Furberg, Martine Kjølberg Moen, Kristin Vardheim Liyanarachi, Åsa Askim, Signe E. Åsberg, Andrew T. DeWan, Tormod Rogne, Gunnar Skov Simonsen, Tom Ivar Lund Nilsen, Bjørn Olav Åsvold, Jan Kristian Damås, Erik Solligård

**Affiliations:** Gemini Center for Sepsis Research at Institute of Circulation and Medical Imaging, NTNU, Norwegian University of Science and Technology, Trondheim, Norway; Clinic of Anesthesia and Intensive Care, St. Olavs hospital, Trondheim University Hospital, Trondheim, Norway; Nord-Trøndelag Hospital Trust, Levanger, Norway; Department of Microbiology and Infection Control, University Hospital of North Norway, Tromsø, Norway; Faculty of Health and Social Sciences, Molde University College, Molde, Norway; Department of Infectious Diseases, St. Olavs hospital, Trondheim University Hospital, Trondheim, Norway; Department of Chronic Disease Epidemiology and Center for Perinatal, Pediatric and Environmental Epidemiology, Yale School of Public Health, New Haven, CT, USA; Research Group for Host-Microbe Interaction, Faculty of Health Sciences, UiT – The Arctic University of Norway, Tromsø, Norway; Norwegian Institute of Public Health, Oslo, Norway; Department of Public Health and Nursing, NTNU, Norwegian University of Science and Technology, Trondheim, Norway; K.G. Jebsen Center for Genetic Epidemiology, Department of Public Health and Nursing, NTNU, Norwegian University of Science and Technology, Trondheim, Norway; Department of Endocrinology, Clinic of Medicine, St. Olavs hospital, Trondheim University Hospital, Trondheim, Norway; Centre of Molecular Inflammation Research, Department of Clinical and Molecular Medicine, NTNU, Norwegian University of Science and Technology, Trondheim, Norway

## Abstract

**Objective:** To examine the effect of sex on risk of bloodstream infections (BSI) and BSI mortality and to assess to what extent known risk factors for BSI mediate this association in the general population.

**Participants:** The prospective, population-based HUNT2 Survey (1995-97) in Norway invited 93,898 inhabitants ≥20 years in the Nord-Trøndelag region, whereof 65,237 (69.5%) participated. 46.8% of the participants were men.

**Exposures:** Sex and potential mediators between sex and BSI; health behaviours (smoking, alcohol consumption), education attainment, cardiovascular risk factors (systolic blood pressure, non-HDL cholesterol, body mass index) and previous or current comorbidities.

**Main outcome measures:** Sex differences in risk of first-time BSI, BSI mortality (death within first 30 days after a BSI), BSI caused by the most frequent bacteria, and the impact of known BSI risk factors as mediators.

**Results:** We documented a first-time BSI for 1,840 (2.9%) participants (51.3% men) during a median follow-up of 14.8 years. Of these, 396 (0.6%) died (56.6% men). Men had 41% higher risk of any first-time BSI (95% confidence interval (CI), 28 to 54%) than women. An estimated 34% of the excess risk of BSI in men was mediated by known BSI risk factors. The hazard ratio (HR) with 95% CI for BSI due to *S. aureus* was 2.09 (1.28 to 2.54), *S. pneumoniae* 1.36 (1.05 to 1.76), and *E. coli* 0.97 (0.84 to 1.13) in men vs women. BSI related mortality was higher in men compared to women with HR 1.87 (1.53 to 2.28).

**Conclusions:** This large population-based study show that men have higher risk of BSI than women. One-third of this effect was mediated by known risk factors for BSI. This raises important questions regarding sex specific approaches to reduce the burden of BSI.

## Introduction

Bloodstream infection (BSI) is a major global burden (1) and may lead to sepsis which constitutes up to 60% of the global mortality burden (2–4). The risk of acquiring BSI depends on the bacterial virulence, host characteristics and biological factors (5–9). Epidemiological studies indicate a male predominance in sepsis, nevertheless former studies on sex differences in incidence and mortality of sepsis have given conflicting results (2, 10–14). Importantly, disparities in immune function between sexes arise from differences in biological characteristics such as anatomy and hormonal status, medical conditions, health behaviours, lifestyle, and exposure to different pathogens (12, 15–17). Most previous studies on sex differences in BSI and sepsis have been performed in small cohorts, mainly from the intensive care unit (ICU), and there are limited population-based studies (12–14, 18, 19). Studies on severe infections and sepsis tend to *adjust* for sex in their analyses (20) without focusing on the mechanisms between sex and infection risk (21).

To assess the impact of sex as a risk factor for BSI we used data from the Norwegian HUNT Study linked with hospital-confirmed clinically relevant BSI to estimate the sex specific risk of any first-time BSI, BSI-related mortality, and first-time BSI caused by the most common infecting bacteria, *Staphylococcus (S*.*) aureus, Streptococcus (S*.*) pneumoniae* and *Escherichia (E*.*) coli*. Further, we aimed to investigate the potential mediating effect of health behaviours, education attainment, cardiovascular risk factors and comorbidities on the associations between sex and BSI.

## Methods

### Study population

The HUNT Study is a large-scale population-based health study conducted in mid-Norway and consists of four consecutive surveys inviting the total adult population approximately every 10^th^ year. The second survey (HUNT2, 1995-97) invited all adult inhabitants ≥20 years (n=93,898) to a clinical examination, non-fasting blood sampling and a comprehensive self-report of health-related topics. Of these, 65,237 (69%) chose to participate. The HUNT study database is regularly updated with information on date of migration and death from the National Registry. More details on the HUNT study are published elsewhere (22). For the purpose of the present study, we excluded 47 (0.07%) participants who had a prior positive blood culture and 1,150 (1.8%) who migrated or died before start of follow-up. A total of 64,040 participants were eligible for analyses (Supplemental Figure 1 Flow-chart).

### Measures

The exposure is sex as registered in the National Registry. The two main outcomes were first-time BSI and BSI related mortality. All BSIs were confirmed at the microbiology laboratories at Levanger, Namsos or at St. Olavs hospitals. The participants were followed-up for incident BSI by a linkage to the Nord-Trøndelag Hospital Trust (HNT HF) sepsis registry ***u***sing the personal identification number of Norwegian citizens (23). We defined BSI mortality as death occurring within 30 days after detection of any BSI. In secondary analyses we assessed first-time BSI caused by the most common bacteria *E. coli, S. aureus* and *S. pneumonia*. Blood cultures solely with microorganisms associated with skin contamination such as coagulase negative *Staphylococcus* species, *Corynebacterium* species and *Cutibacterium* species were not considered as BSI (24).

Mediators are variables that are causally located between exposure and outcome variables, and that partly explain the effect of exposure on outcome (25). We used three distinct sets of mediators measured at inclusion; 1) health behaviours (smoking and alcohol use) and educational attainment; 2) cardiovascular risk factors (body mass index (BMI, kg/m^2^), systolic blood pressure (mmHg) and non-high-density lipoprotein cholesterol (non-HDL, mmol/L); 3) comorbidities defined by self-report of cardiovascular disease (history of myocardial infarction, angina pectoris, and/or stroke), diabetes, cancer history, lung disease (asthma or chronic obstructive pulmonary disease) and standardised measurements of kidney disease (estimated glomerular filtration rate (eGFR) < 60 ml/min/1.73 m^2^) (22). The three sets of mediators reflect known risk factors for BSI. Details about the measurements and categorisation of mediators are included in the supplemental material.

### Statistical analyses

We used Cox proportional hazard regression analysis to estimate the hazard ratios (HRs) with 95% confidence intervals (CIs) of a first-time BSI and BSI mortality in men compared to women. Attained age was used as the time scale. Start of follow-up was defined by the availability of data in the sepsis registry. For residents belonging to Levanger hospital from 1 January 1995, and for residents belonging to Namsos hospital from 1 September 1999. Patients referred to St. Olavs hospital, the tertiary referral centre, BSI information was included depending on their primary hospital. Participants contributed person-years from inclusion date in HUNT2 except for participants having Namsos as their primary hospital, they entered the HNT HF sepsis registry from 1 September 1999. Data from all hospitals were available through 2011 (23).

In the analysis of BSI risk, participants were followed until their first BSI, for BSI-mortality participants were followed until death within 30 days of any BSI episode, and for both analyses participants were followed until migration out of Nord-Trøndelag, death of all causes or end of follow-up set to 31 December 2011, whichever occurred first. The proportional hazards assumption was examined by visual inspection of log-log plots and tests of Schoenfeld residuals. Using Stata *stcompadj*, we estimated cumulative incidence and mortality from start of follow-up to first-time BSI and BSI mortality, accounting for death by all causes as a competing risk.

In secondary analyses we estimated HRs and cumulative incidence of first-time BSI caused by the most common infecting bacteria; *E. coli, S. aureus* and *S. pneumoniae* using Cox regression and cumulative incidence. *To address whether menopause affects women’s risk of BSI we conducted a sensitivity analysis stratified by age below and above 50 years*.

The proportion of excess risk of BSI in men mediated by known BSI risk factors was estimated using an *inverse odds weighting (IOW)* procedure (26, 27). IOW is a counterfactual method that enables a decomposition of the *total effect* of the exposure (sex) on the outcome (first-time BSI) into a *natural direct effect (NDE)* from exposure on outcome, and a *natural indirect effect (NIE)* through multiple mediators jointly fitted in the model (25, 26). The method is robust to unmeasured common causes of the mediators (25).

The inverse odds weights were obtained by regressing the exposure (sex) on all mediators of interest with age as a covariate. In our analysis, the total effect is best interpreted as the total association between exposure (sex) and outcome (first-time BSI), the natural indirect effect is the proportion of excess BSI risk in men mediated by conventional BSI risk factors, whereas the natural direct effect is the proportion of excess BSI risk in men not associated with these factors. The proportion mediated is the percent of the total association that is mediated through the conventional risk factors. A graphic description of the mediation analyses is given in Figure 1. We did not estimate the NIE of individual mediators separately as it may not be appropriate when the mediators affect each other or when single mediator-outcome confounders may be affected by exposure (25). Instead, we estimated the NIE with a sequential mediation approach using three models. This approach assumes that the cardiovascular risk factors and further the comorbidities can be causal descendants of the health behaviours and educational attainment (25, 28). In model 1, we assessed health behaviours and education; in model 2, we added BMI, systolic blood pressure and non-HDL cholesterol; and in model 3, comorbidities were included to the complete set of mediators. The sequential approach further implies that model 3 reflects the best interpretation of the mediation analyses as all mediators and age are included (25).

**Figure 1.**
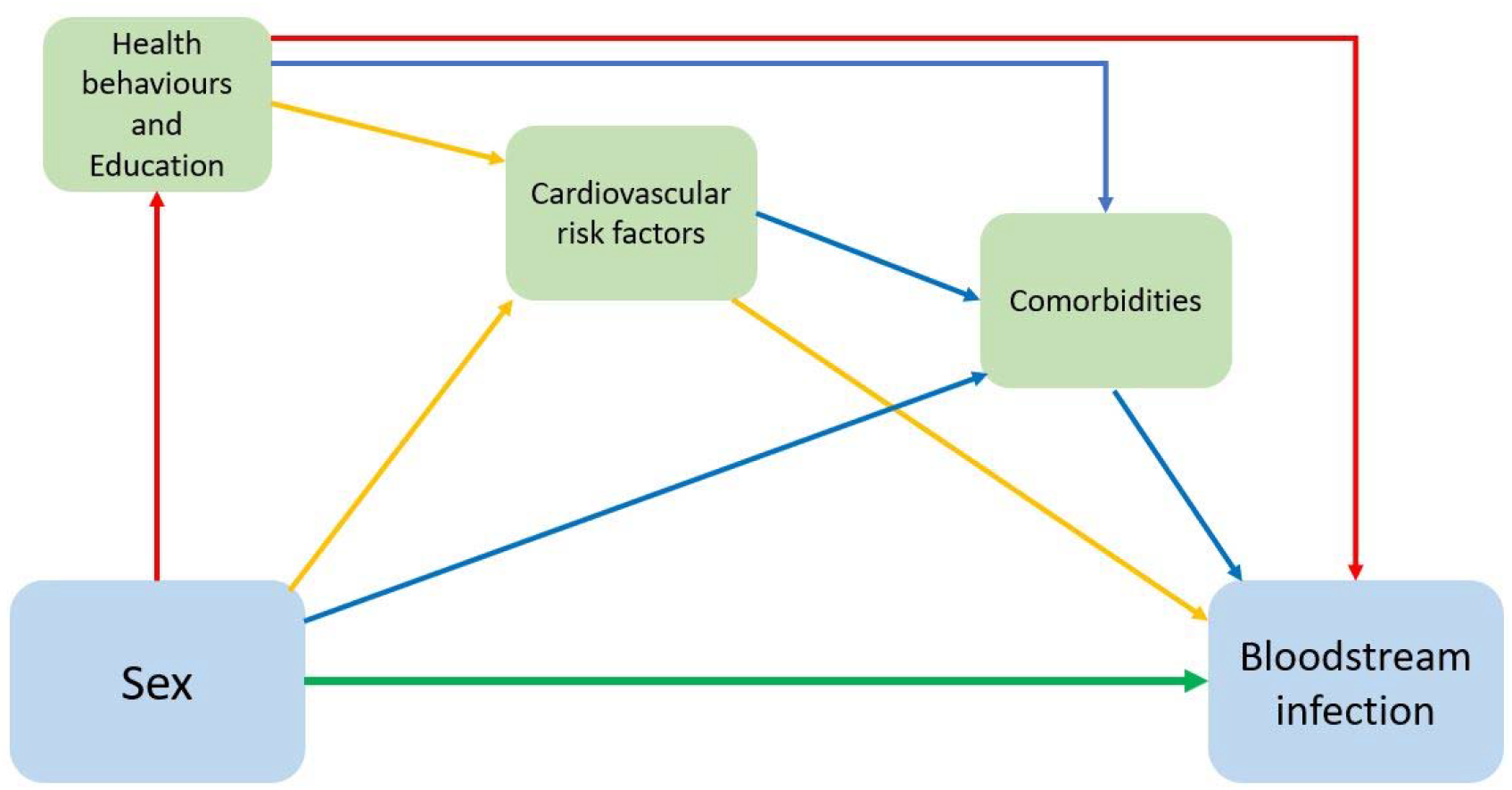
Mediation analysis. Diagram of the direct and indirect (i.e. mediated) effects of **sex** on **bloodstream infection**. The green arrow represents the natural direct effect of the association. Red arrows represent model 1, proportion mediated by health behaviours and education attainment. Yellow arrows represent model 2, proportion mediated jointly by health behaviours, education, and cardiovascular risk factors. Blue arrows represent model 3, proportion mediated jointly by health behaviours, education, cardiovascular risk factors and comorbidities. Model 1) Smoking, alcohol use and educational attainment at baseline. Model 2) Systolic blood pressure, non-high-density lipoprotein cholesterol and Body Mass index. Model 3) Cardiovascular disease, chronic kidney disease, diabetes, history of cancer, and chronic lung disease.

We performed bootstrapping based on 1,000 replications to derive percentile-based CIs for all mediation parameters (28). We present the NDE and NIE as HRs with 95 % CIs. The proportion mediated on the log scale was calculated using the formula (lnHR_NIE_ /lnHR_TOTAL_) (26). All statistical analyses were performed using Stata version 16.0. A detailed description of the steps in the IOW analyses is included in supplemental material.

### Ethics

The study was approved by the Regional Committee for Medical and Health Research Ethics of Central Norway (REK no 2012/153 and REK no 94135), and by the HUNT data access committee. Participation in HUNT2 was voluntary, and participants signed a written consent to data collection and linking their data to other registers.

## Results

### Population characteristics

Among 64,040 participants, of whom 46.8% were men, 1,840 (2.9%) experienced a BSI and 396 (0.6%) died within 30 days after an episode of BSI. The median age at inclusion was similar for both sexes. 51.3% of first-time BSI cases were men, and 56.6% of BSI-related deaths occurred in men (Table 1). Both men and women who experienced a BSI were older (median age at inclusion 67.4 and 68.0 respectively), and they had a higher comorbidity burden than participants who did not have a BSI during follow-up.

**Table 1:** Baseline characteristics of the study population at inclusion in HUNT2, n=64,040.

|  | Men | Women |
| --- | --- | --- |
| Total population n (%) | 29962 (46.8) | 34087 (53.2) |
| First-time BSI n (%)<sup>1</sup> | 943 (51.3) | 897 (48.7) |
| BSI mortality<sup>2</sup> n (%) | 224 (56.6) | 172 (43.4) |
| Age (mean, IQR) | 48.6 (36.5-62.9) | 48.7 (36.2-64.2) |
| Smoking |  |  |
| Current | 8334 | 9726 |
| Prior | 9422 | 6516 |
| Never | 10668 | 15230 |
| Alcohol use |  |  |
| < 1 unit/ 2 weeks | 8448 | 16069 |
| 1-7 units/ 2 weeks | 14258 | 14484 |
| 8-14 units/ 2 weeks | 4602 | 1861 |
| ≥ 15 units/ 2 weeks | 1643 | 272 |
| Education |  |  |
| < 10 years | 20625 | 22259 |
| 10-12 years | 2302 | 3436 |
| > 12 years | 5650 | 6412 |
| BMI (kg/m<sup>2</sup>) |  |  |
| <18.5 | 118 | 349 |
| 18.5-24.9 | 10498 | 14736 |
| 25-29.9 | 14757 | 12345 |
| 30-34.9 | 3674 | 4640 |
| 35-39.9 | 496 | 1236 |
| ≥ 40 | 74 | 344 |
| Systolic blood pressure (mmHg) | 137 | 131 |
| median (IQR) | (127-150) | (118-149) |
| Non-HDL cholesterol (mmol/L) | 4.5 | 4.3 |
| median (IQR) | (3.7-5.3) | (3.5-5.3) |
| Comorbidities |  |  |
| Cardiovascular disease<sup>3</sup> | 2918 | 2014 |
| Chronic kidney disease | 979 | 1802 |
| Diabetes | 970 | 895 |
| Cancer history | 878 | 1413 |
| Chronic lung disease<sup>4</sup> | 1183 | 1011 |

BSI= Bloodstream infection, n= Numbers, IQR= Interquartile range, BMI= Body mass index. HDL= High-density lipoprotein
1) Percentage of total first-time BSI in both sexes.
2) BSI mortality was defined as all-cause mortality within 30 days after a BSI.
3) History of myocardial infarction, angina pectoris and/or stroke.
4) History of chronic obstructive pulmonary disease or asthma.

### Risks of BSI and BSI mortality

Men were more likely to experience a first-time BSI and to die from a BSI compared to women. After adjustments for age, men had 1.41 (HR, 95% CI 1.28-1.54) higher risk of first-time BSI, and 1.87 (HR, 95% CI 1.53 – 2.28) higher risk of dying from a BSI within 30 days (Table 2). In analyses by the most common infecting bacteria compared to women, men had a 2.09-fold (HR, 95% CI 1.28 to 2.54) increased risk of BSI caused by *S. aureus*, and 1.36 (HR, 95% CI 1.05 to 1.76) increased risk of *S. pneumonia*. The corresponding result for *E. coli* did not show higher risk in men with HR of 0.97 (95% CI 0.84 to 1.13) (Table 3).

**Table 2.**
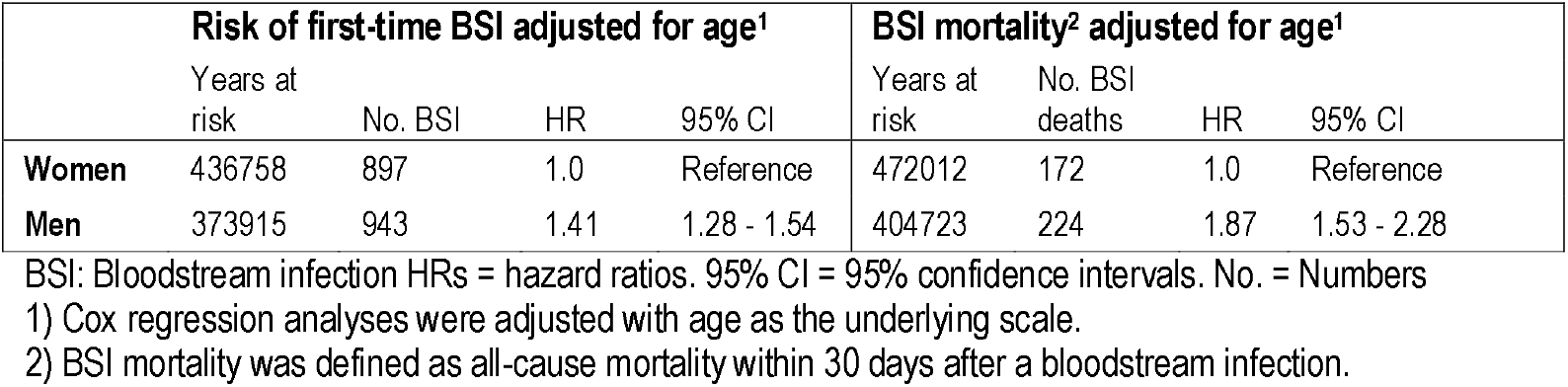
Associations of sex with risk of bloodstream infection and BSI mortality.

**Table 3.** Associations of sex with risk of bloodstream infections caused by the most common bacteria.

|  | Risk of *S. aureus* BSI<sup>1</sup> |  |  |  | Risk of *S. pneumoniae* BSI<sup>1</sup> |  |  | Risk of *E. coli* BSI<sup>1</sup> |  |  |
| --- | --- | --- | --- | --- | --- | --- | --- | --- | --- | --- |
|  | Years at risk | No. BSI | HR | 95% CI | No. BSI | HR | 95% CI | No. BSI | HR | 95% CI |
| **Women** | 436758 | 83 | 1.0 | Reference | 113 | 1.0 | Reference | 399 | 1.0 | Reference |
| **Men** | 373915 | 129 | 2.09 | 1.28 - 2.54 | 119 | 1.36 | 1.05 - 1.76 | 285 | 0.97 | 0.84 - 1.13 |

BSI: Bloodstream infection HRs = hazard ratios. 95% CI = 95% confidence intervals. No. = Numbers
1) Cox regression analyses were adjusted for age as the underlying scale.

Figure 2 illustrates the above findings by graphing the age-adjusted cumulative incidence of first-time BSI and cumulative BSI mortality. The cumulative incidence of BSI was higher among men than women after the first five years of follow-up. For BSI mortality the sex difference was apparent after the first 2.5 years of follow-up and during follow-up the sex differences in mortality increases substantially. We present cumulative incidence curves for the three most common infecting bacteria in Figure 3A, B and C. Men had higher incidence of *S. aureus*, especially after the first 7 years of follow-up, whereas *E. coli* had higher cumulative incidence among women.

**Figure 2:**
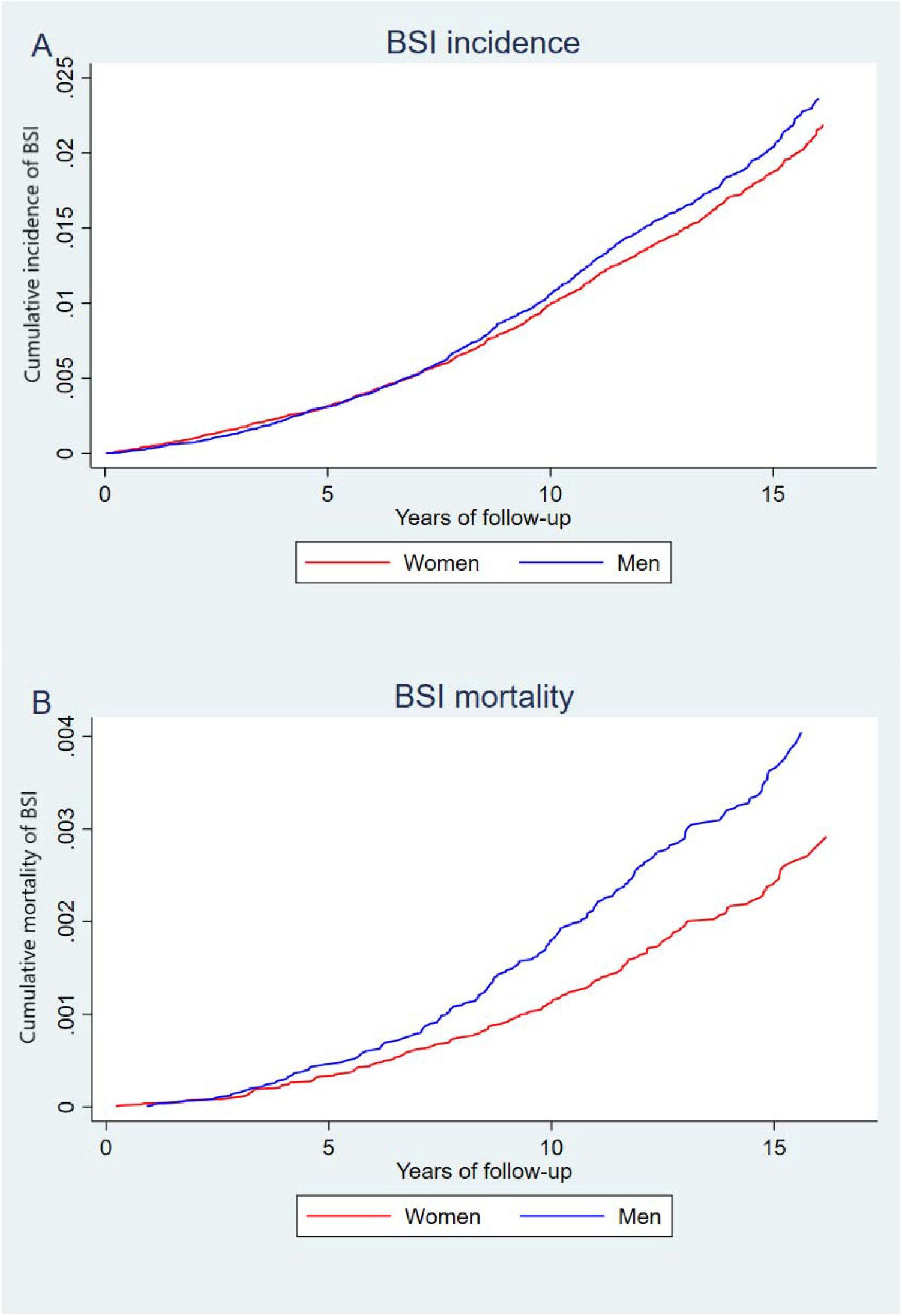
Sex differences in cumulative incidence and mortality of BSI. Age-adjusted sex difference in cumulative incidence of BSI (A), and in cumulative mortality (B), estimated for age 49.99 (the mean age of the total population). Note: due to the variation in incidence of different outcomes the scale of the Y-axis is not uniform across the panels.

**Figure 3:**
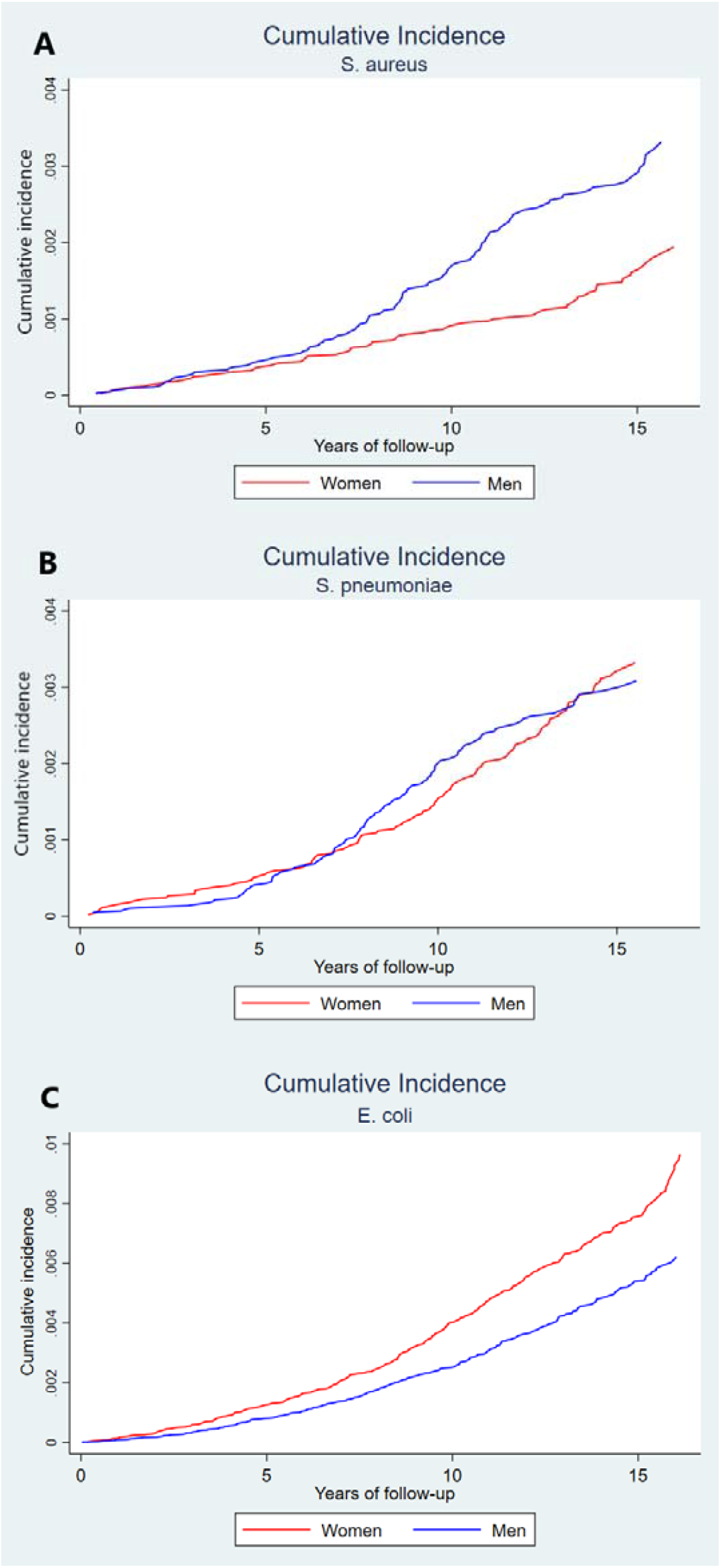
Sex differences in cumulative incidence of BSI caused by the most common bacteria. Age-adjusted sex difference in cumulative incidence of *S. aureus* (A), *S. pneumoniae* (B), and *E. coli* (C), estimated for age 49.99 (the mean age of the total population). Note: due to the variation in incidence of different bacteria the scale of the Y-axis is not uniform across the panels.

The age-stratified analyses at estimated menopause (≤50 years) did not reveal any clear sex difference in risk of BSI before menopause. However, the youngest age-group had few episodes of BSI, and few persons died from a BSI (Supplemental Table 1).

### Mediation analyses

In Table 4 we present the total effect, the natural direct and indirect effects of sex on risk of first-time BSI. Compared with women men had an estimated HR of 1.40 (95% CI 1.24 to 1.55) for first-time BSI. Behavioural risk factors and education mediated 10% of the total effect (model 1), adding the cardiovascular risk factors mediated 5% (model 2), whereas the whole set of mediators, including comorbidities, jointly mediated 34% of the total effect (model 3). The mediation results indicate that male sex is a risk factor for BSI, but also that men with increased burden of conventional BSI risk factors are at particular risk.

**Table 4:** **Mediation of the associations between sex and BSI by behavioral risk factors, educational attainment, cardiovascular risk factors and comorbidities**.

|  | Risk of first-time BSI |
| --- | --- |
| **Model 1** |  |
| **Mediation by behavioural riskfactors<sup>1)</sup> and education** | **HRs (95% CI)<sup>a)</sup> <sup>b)</sup>** |
| Total effect | 1.40 (1.24 – 1.55) |
| Natural direct effect (NDE) | 1.36 (1.18 – 1.57) |
| Natural indirect effect (NIE) | 1.04 (0.97 – 1.07) |
| Proportion mediated<sup>c)</sup> | 10 % |
| **Model 2** |  |
| **Mediation by behavioural risk factors<sup>1)</sup>, education, and cardiovascular risk factors<sup>2)</sup>** | **HRs (95% CI)<sup>a)</sup> <sup>b)</sup>** |
| Total effect | 1.40 (1.24 – 1.55) |
| Natural direct effect (NDE) | 1.38 (1.19 – 1.58) |
| Natural indirect effect (NIE) | 1.02 (0.92 – 1.07) |
| Proportion mediated<sup>c)</sup> | 5 % |
| **Model 3** |  |
| **Mediation by behavioural risk factors<sup>1)</sup>, education, cardiovascular risk factors<sup>2)</sup> and comorbidity risk factors<sup>3)</sup>** | **HRs (95% CI)<sup>a)</sup> <sup>b)</sup>** |
| Total effect | 1.40 (1.24 – 1.55) |
| Natural direct effect (NDE) | 1.25 (1.05 – 1.47) |
| Natural indirect effect (NIE) | 1.12 (1.02 – 1.17) |
| Proportion mediated<sup>c)</sup> | 34 % |

BSI: Bloodstream infection HRs = hazard ratios. 95% CI = 95% confidence intervals.
1) Smoking, alcohol use and educational attainment at baseline.
2) Systolic blood pressure, non-high-density lipoprotein cholesterol and Body Mass Index.
3) Cardiovascular disease, chronic kidney disease, diabetes, history of cancer, or chronic lung disease.
a) Percentile-based bootstrap CIs are reported.
b) Estimates are adjusted for age as a covariate.
c) Proportion mediated: $(\ln \text{HR}_{\text{NIE}} / \ln \text{HR}_{\text{TOTAL}})$ .

## Discussion

In this large Norwegian population-based study with a follow-up of more than 15 years, male sex was associated with 1.41 times higher risk of BSI and 1.87 times higher risk of dying from a BSI. An estimated 34% of the increased risk of BSI in men was mediated by conventional BSI risk factors. We additionally found that men had 2.09 times higher risk of BSI caused by *S. aureus* compared to women. Importantly, our findings indicate that male sex contributes as an individual and important risk factor for BSI. However, they do not refute the impact of baseline burden of health behaviours, education and pre-existing chronic health conditions in sex disparities seen in BSI.

### Comparison with other studies

There are few population-based studies on sex differences in the epidemiology of BSI and to our knowledge, no previous mediation analysis has combined conventional risk factors with validated BSI events to examine this topic. The rich baseline information from HUNT2 including chronic medical conditions, cardiovascular risk factors, health behaviours and education attainment allowed us to investigate how these factors mediated the association between sex and BSI. We used the IOW approach which is known to be robust using multiple mediators en bloc, and allowed us to implement causal mediation analyses in a time-to-event context (25, 26). The sequential approach enabled us to examine the portion mediated through health behaviours and education, which accounted for 10 %. Adding cardiovascular risk factors the proportion mediated accounted for 5%, indicating some interactions or common pathways for the mediators (25). The complete model accommodate the assumptions required, and most likely reflects the best modelling of the associations (25). In our analyses the proportion mediated by age and all mediators combined was 34%. Interventions to reduce modifiable risk factors in the population will likely reduce the burden of BSI, particularly in aging men with high burden of known risk factors. This is an important finding that deserves further investigations.

The population-based design ensures that all BSI occurring in residents of a defined geographical area are included, which is an advantage over ICU cohorts (29). Our results are supported by one population-based BSI study including 1,051 patients with positive blood cultures showing that men had higher risk of BSI. In particular they found that BSI incidence increases sharply by age, and men had twice the rate of *S. aureus* BSI (18). Another population-based study from Canada comprising 9,266 patients with BSI admitted to ICU found that male sex is a risk factor for BSI (19). From the same research group, a recent study restricted to persons aged ≥65, found that men were at increased risk compared to females (incidence rate ratio 1.44, 95% CI 1.32-1.59) and the sex difference was most pronounced in the oldest patients (30). On the other hand, the global burden of disease study found that age-standardised sepsis incidence was higher among women, while sepsis-related mortality was higher among men (2). This study included results from 195 countries and comprised all age groups. They found a substantially higher sepsis incidence in low-income countries, and the pattern of sepsis incidence and mortality varied according to location, which is not directly comparable to our study population.

We identified higher BSI mortality in men which is in line a population-based study of BSI in critically ill adults (31), and a recent study on infection related death in UKBiobank (32). Conversely, some ICU studies report higher sepsis-related mortality in women. A retrospective study with 18,757 adult ICU patients, found higher mortality in women (13). The same pattern was observed in a Swedish study on community acquired sepsis where women had 28% higher odds of dying (10), and in a smaller ICU cohort from Germany studying 709 adults (11). This conflicting evidence concerning sex differences in mortality is most likely due to the heterogeneity of BSI and sepsis depending on the aetiology and the cohort studied (29, 31), but may also be affected by sex differences in immune responses (15–17, 33) and differences in treatment (13, 34).

The second most common infecting agent in our study was *S. aureus* which was far more common in men. *S. aureus* is associated with superficial infections of soft tissues with the potential for severe invasive infections and is the most important cause of BSI-associated death (35, 36). Previous studies indicate higher probability of nasal colonization in men which is a major risk factor for invasive *S. aureus* infections (37). Other studies show that circulating levels of testosterone and use of hormone contraceptives among females alter nasal colonization indicating that sex hormones affect the immune response to *S. aureus* (38, 39). The substantially higher prevalence of *S. aureus* colonization in men is of particular interest, and preventive measures like eradication or temporary suppression could lower the risk of invasive infections (40), which is especially important in hospitalized patients.

In a sensitivity analysis we found that the sex differences in BSI risk are evident after predicted age of menopause indicating that alterations in both innate and adaptive immune functions with age may be sex specific. Aging is associated with chronic inflammation and a generally reduced immune function. Women face an abrupt decline during menopause, whereas men have a steady decline from second decade of life (41). As in former studies, our study points out that advancing age is a risk factor for developing and dying from BSI and elderly men are at particular risk (18, 19, 30).

### Strengths and limitations of the study

Major strengths of our study include its large size, the population-based design, long-term follow-up and linkage to all microbiological records which represent the gold standard to identify BSI (29). Reviews of medical records of patients with *S. aureus* and *S. pneumoniae* BSI in this cohort showed that ∼98% met the 2001 sepsis criteria (35, 42). Our definition of BSI as a laboratory verified positive blood culture, excluding blood cultures solely with microorganisms associated with skin contamination, ascertains the accuracy of the outcome studied. We were able to study BSI incidence and BSI mortality in a large population without the potential referral bias seen in single institution studies of BSI. The complete ascertainment of all BSI allows for an accurate estimation of incidence and mortality in the population, with the potential of risk factor identification and mediation analyses.

There are some limitations of our study that merit attention. First, we lack information on immunosuppressive medication and use of statins before or at time of infection which are known risk factors for BSI and BSI mortality (9, 43). Second, investigating BSI is dependent on clinician’s suspicion and decision to submit blood cultures for testing, with the chance of some undetected cases. Further, we cannot rule out if the clinical presentation of infections is different in men and women and that this could result in disproportionate blood culture sampling depending on sex. Third, the mediators are only measured once at inclusion to HUNT2 and could have changed during the 15 years of follow-up. This potential misclassification would most likely be non-differential and lead to underestimation of the mediating effect (44). Another concern is the subjective assessment of some mediators, and dichotomised mediators are more prone to possible mediator misclassification. This could lead to underestimation of the indirect effect and overestimation of the direct effect (45).

Despite these limitations and to the best of our knowledge, we present the first large, epidemiological study elucidating sex differences in BSI. With a non-selected, homogenous population-based study with long follow-up on blood-culture verified BSI with known pathogen, our study contributes to a growing body of evidence that sex influence BSI risk.

### Meaning of the study and future research

This study provides important information for clinicians, researchers and policymakers concerning BSIs. We show that sex disparities in BSI cannot be explained by comorbidity burden, health behaviours and educational level alone. The clinical implications of our findings largely depend on elucidation of the underlying mechanisms behind the observed sex differences.

Sex affects the shape of immune responses which is attributed to genetic, hormonal and environmental factors (15, 16, 33). The human X-chromosome encodes a number of critical genes involved in the regulation of immune functions (33). It is increasingly clear that sex extensively influences the host immune responses, but this sexual dimorphism is underappreciated, and sex bias is a major challenge in clinical trials (20, 34, 46). Sex hormones act as important modulators of immune functions and responses; testosterone and progesterone are immunosuppressive, while oestradiol is immunoenhancing (16). Few human studies have investigated sex hormones’ effects in sepsis. Interestingly, in covid-19 where men are more prone to a severe course, the use of antiandrogens in men have shown promising results on severity (47). Furthermore, in a review of health records in post-menopausal women with regular use of oestradiol, the fatality risk of covid-19 is reduced by more than 50% (48).

Future perspectives of our results include the need for targeted research on how these sex differences could be addressed to achieve a longer and healthier life for both men and women. Additional work should focus on how genetics and epigenetics as well as sex hormones play a role in the sex disparities seen in infectious diseases. Knowledge of mediating factors together with recognition of sex differences in severe infections are important for public health leaders, researchers and clinicians as it can inform preventive actions and identify individuals especially at risk (21, 49).

## Conclusion

In this large population-based cohort, men had an increased risk of BSI and BSI mortality, where 34% risk of BSI was explained by known risk factors. BSI represents an important global burden of disease, and by establishing the presence of sex differences in BSI our study provides as a catalyst for additional investigations. This will advance our understanding of sex specific host-pathogen interactions and sex hormone’s immune actions, and potentially lead to targeted management strategies to prevent BSI and sepsis in both men and women.

## Data Availability

The data used in this work is available by application from HUNT. https://hunt-db.medisin.ntnu.no/hunt-db/

## Acknowledgements

The Trøndelag Health Study (the HUNT study) is a collaboration of the HUNT research center (Faculty of Medicine and Health Sciences, NTNU, Norwegian University of Science and Technology), Trøndelag County Council, Central Norway Regional Health Authority, and the Norwegian Institute of Public Health.

We sincerely thank Arne Mehl (1948-2021) for his extensive and thorough work with the HNT HF sepsis registry, the microbiology departments of Levanger, Namsos and St. Olav hospitals for providing microbial data, and the Department for Research at Nord-Trøndelag Hospital Trust for assistance with data linkage.

## Contribution

RMM, LG, MKM, KVL, SEÅ, TN, JKD and ES conceived and designed the study. TN, BOÅ, LG, ATD, TR, JKD and ES supervised the study. RMM, BOÅ, ÅA and LG contributed in the acquisition of data for statistical analyses. RMM analyzed the data, prepared tables and figures. RMM, LG, ASF, MKM, KVL, ÅS, SEÅ, GSS, TR, TN, ATD, BOÅ, ES and JKD interpreted the results and drafted the manuscript. RMM, LG, ASF, MKM, KVL, ÅA, SEÅ, ATD, TR, GSS, TN, BOÅ, JKD and ES contributed to the discussions and revised the manuscript critically for important intellectual content. All the authors have read and approved the final version of manuscript and agreed to be accountable for all aspects of the work.

