## Supplementary material for "Sex differences in incidence and mortality of bloodstream infections. Results from the population-based HUNT study in Norway"

Randi Marie Mohus et al.

**Supplementary method:** Study population and mediators.

**Supplementary table 1:** Procedure of estimating mediation parameters using the IOW method.

**Supplementary table 2:** Age-stratified associations of sex with risk of bloodstream infection.

**Supplementary figure:** Flow Chart of Study Recruitment and Follow-up

Supplementary methods:

Study population

The HUNT study database is regularly updated with information on date of migration and death from the National Registry. The Nord-Trøndelag region in Norway has a population of 130,000, where approximately 70% is served by Levanger hospital and 30% is served by Namsos hospital. The tertiary referral centre is St. Olavs hospital in Trondheim. The population is stable with a net out-migration of 0.3% per year, and ethnically homogeneous (97% Caucasians) [1].

The Nord-Trøndelag Hospital Trust (HNT HF) sepsis registry has prospectively recorded information on all clinically relevant BSI events at Levanger hospital from 1 January 1995, and Namsos hospital from 1 September 1999. All HUNT2 participants with a positive blood culture recorded at St. Olavs hospital were included in the registry from 1 January 1995.

Mediators

The creatinine values used in the estimated glomerular filtration rate (eGFR) calculation were measured in non-fasting serum blood samples drawn by trained nurses and analyzed at the Central Laboratory at Levanger Hospital. eGFR was estimated from recalibrated creatinine values using the Modification of Diet in Renal Disease (MDRD)-formula [2]. Body mass index (BMI) was calculated as weight (kg) divided by the squared value of height (m^2^), measured by trained nurses at the clinical examination at inclusion in HUNT2 with the participants wearing light clothing and no shoes. BMI was categorized as recommended by WHO (<18.5, 18,5-24.9, 25-29.9, 30-34.9, 35.0 -39.9, ≥ 40 kg/m^2^). Systolic blood pressure was measured three times at 1-minute intervals using an automatic oscillometric method (Dinamap, Critikon, Florida, USA) after a person had come to rest, with cuff size adjusted to arm circumference. We used the mean of the second and third measurments. Serum total and high-density lipoprotein (HDL) cholesterol were analysed using enzymatic colorimetric methods (Boeheringer Mannheim, Germany). Non-HDL cholesterol was calculated as the difference between total and HDL cholesterol.

Smoking was defined from several questions on past and current smoking; as “current smoking” (smoking tobacco daily), “prior smoking” (any prior daily tobacco smoking) or “never smoked”. Alcohol use as “never drink alcohol”, “1-7 units of alcohol in two weeks”, “8-12 units alcohol in two weeks” or “more than 15 units in two weeks”. Educational attainment was categorized as <10 years, 10-12 years and >12 years of schooling.

Supplementary Table 1: Procedure of estimating mediation parameters using the IOW method [3, 4]

| Steps | Procedure | Stata code |
| --- | --- | --- |
|  | Preparing the data | *User written program to estimate mediation parameters  Capture program drop IOW  Program IOW, rclass  Capture drop predprob inverseodds wt_iow |
| Step 1: Exposure model | The exposure model is run by regressing the exposure on all mediators and age as a covariate using logistic regression | *model 1  logit Sex i. smoking i. alc i. edu_cat age  *model 2  logit Sex i. smoking i. alc i. edu_cat systBP non_HDL i2. BMI age  *model 3  logit Sex i. smoking i. alc i. edu_cat systBP non_HDL i2. BMI i. LungDis i. CardDis i. diabetes i. cancer i. RenalDis age |
| Step 2: Create inverse odds weights | Based on the logistic regression models in step 1, the inverse odds weights are created by estimating the inverse of the predicted odds for each observation in the exposed group. The exposed and unexposed groups are then reweighted as follows: exposed = inverse odds, unexposed = 1 | *obtain predicted probability for each individual based on the above regression models:  predict predprob, p  *calculate each individual’s inverse odds from the predicted probability:  gen inverse odds = ((1-preprob)/predprob)  gen wt_iow = 1 if sex==0  replace wt_iow = inverseodds if sex==1 |
| Step 3: Total effect model | The total effect of the exposure is estimated by using Cox regression model | stset eof, id(PID) failure(bacteriemia) origin(birthyear) enter(Enterdate) scale(365.25)  stcox sex  matrix bb_total = e(b)  scalar b_total =bb_total [1,1]  return scalar b_total=bb_total [1,1] |
| Step 4: Natural direct effect model | The direct effect model is similar to the total effect model, but includes the inverse odds weight constructed from the mediators, instead of controlling for the mediators themselves. | *Estimate the direct effect of sex on BSI by means of a weighted Cox proportional hazards model with the weights (pweight=wt_iow) achieved in step 2:  stset eof [pweight=wt_iow], id(PID) failure(bacteriemia) origin(birthyear) enter(Enterdate) scale(365.25)  stcox sex  matrix bb_direct = e(b)  scalar b_direct = bb_direct [1,1]  return scalar b_direct = bb_direct [1,1] |
| Step 5: Natural indirect effect model | The indirect effect is estimated by subtracting the direct effect from the total effect. | return scalar b_indirect=b-total-b_direct |
| Step 6: Proportion mediated | The proportion mediated is estimated using the formula: lnHR_NIE_ /lnHR_TOTAL_ | return scalar b_mediated=((b_indirect)/b_total)  end |
| Step 7: Estimate standard errors | The standard errors and confidence intervals are estimated by bootstrapping. | bootstrap r(b_indirect) r(b_direct) r(b_total) r(b_mediated), seed (32222) reps(1000):IOW  estat bootstrap, all |

Supplementary Table 2: Age^1^-stratified associations of sex with risk of bloodstream infection and mortality

|  | **Risk of first-time BSI** | | | | **BSI mortality^2^** | | | |
| --- | --- | --- | --- | --- | --- | --- | --- | --- |
|  | Years  at risk | No. BSI | HR | 95% CI | Years  at risk | No. deaths | HR | 95% CI |
| **Women** 50 years and younger | 244845 | 163 | 1.00 | Reference | 245741 | 15 | 1.00 | Reference |
| **Men** 50 years and younger | 216548 | 145 | 0.99 | 0.79 – 1.23 | 217181 | 18 | 1.32 | 0.66 -2.61 |
| **Women** older than 50 years | 191913 | 734 | 1.00 | Reference | 194412 | 157 | 1.00 | Reference |
| **Men** older than 50 years | 157367 | 798 | 1.51 | 1.36 -1.67 | 159761 | 206 | 1.92 | 1.56 – 2.37 |

BSI: Bloodstream infection HRs = hazard ratios. 95% CI = 95% confidence intervals. No. = Numbers

1) Age at inclusion in HUNT2.

2) BSI mortality was defined as all-cause mortality within 30 days after a bloodstream infection.

Supplementary Figure: ***Flow Chart of Study Recruitment and Follow-up***

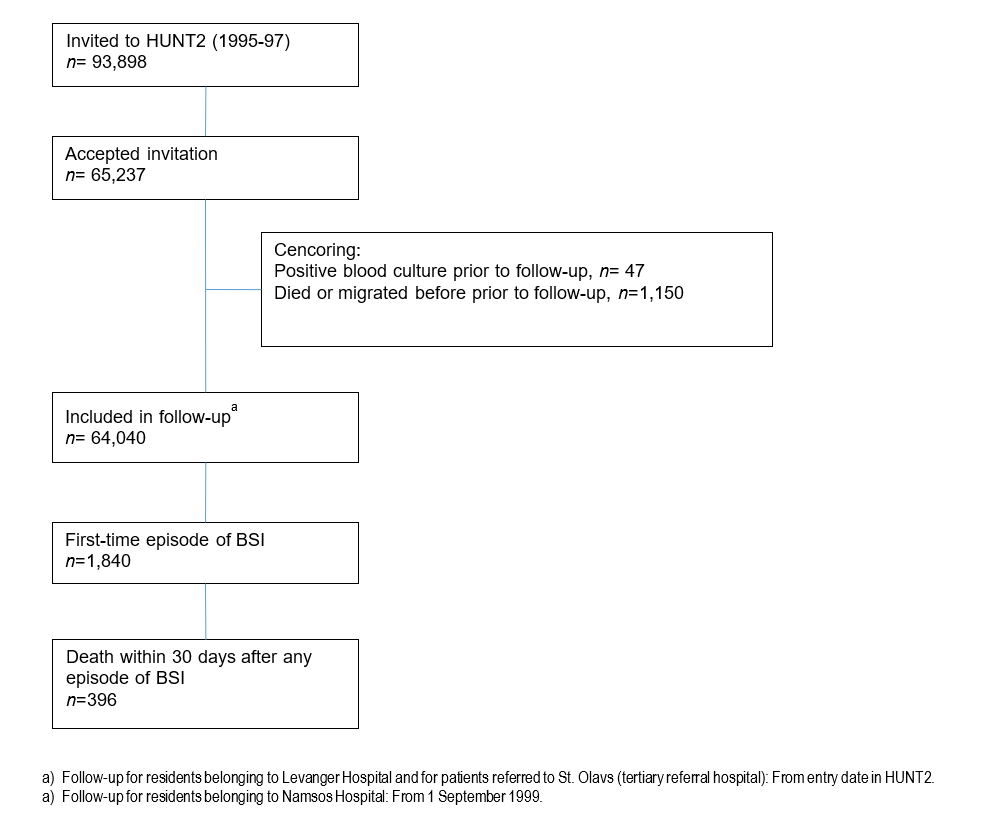
